# An Assessment of Bayesian Model-Averaged Logistic Regression for Intensive-Care Prognosis

**DOI:** 10.1101/2020.06.08.20124412

**Authors:** Richard Dybowski

## Abstract

Logistic regression is the standard method for developing prognostic models for intensive care, but this approach does not take into account the uncertainty in the model selected and the uncertainty in its regression coefficients. This weakness can be addressed by adopting a Bayesian model-averaged approach to logistic regression; however, with respect to the dataset used for our study, we found maximum likelihood to be as effective as the more elaborate Bayesian approach, and an implementation of model averaging did not improve performance. Nevertheless, the Bayesian approach has the theoretical advantage that it can exploit prior knowledge about regression coefficient and model probabilities.

## 1. INTRODUCTION

The primary role of intensive care units (ICUs) is to monitor and stabilize the vital functions of patients with life-threatening conditions. In order to aid ICU nurses and intensivists with this work, *scoring systems* have been developed to express the overall state of an ICU patient as a numerical value. In 1981, Knaus et al^1^ proposed an index of patient severity called APACHE (APACHE I) for use within ICUs, the value of APACHE increasing as the state of a patient declines. The APACHE I score is an additive model based on demographic and physiological attributes, such as age and serum bilirubin:

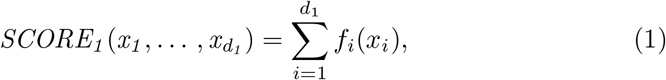

where function *f*_*i*_(·) gives the number of points associated with attribute value *x*_*i*_. For physiological attributes, *f*_*i*_(*x*_*i*_) increases from zero as the divergence of *x*_*i*_ from clinical normality increases.

APACHE I was superseded by SAPS I^2^ in 1984 and APACHE II^3^ in 1985, but, in all three cases, attribute selection and functional form for (1) were determined subjectively through panels of experts. Nevertheless, in spite of the subjectivity of (1), a number of intensivists (e.g., Chang et al^4^) have used this type of score to estimate probabilities of a defined outcome (e.g., alive whilst in hospital) through logistic regression:

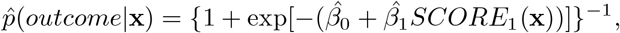

where 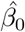 and 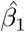are regression coefficients, and x is the vector of values 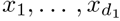.

In 1985, Lemeshow et al^5^ replaced (1) with the linear combination

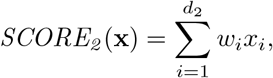

in which weights 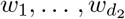 were obtained objectively as logistic regression coefficients:

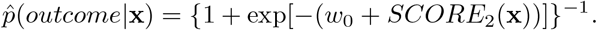

This objective approach was used in the development of a number of scoring systems, including APACHE III,^6^ SAPS II^7^ and MPM II.^8^ A comparison of scoring systems has shown that those derived by logistic regression perform similarly to each other but are better than those obtained subjectively.^9^

Prognostic logistic regression models have been developed within intensivecare medicine for a number of reasons:

- Conditional probabilities of outcome can be used to stratify patients at the outset of a therapeutic drug trial.^10^ This is done to exclude those patients unlikely to display a benefit because their probability of mortality is either too low or too high.
- Outcome models have been used to compare different ICUs,^11^ but such comparisons must be treated with caution.^12^ They can also be used to provide a baseline to assess how a change of policy within a single ICU affects patient outcome.
- A potential (albeit controversial) use of prognostic models is as an aid to the identification of those cases unlikely to benefit from continued care.^13^
- All the models developed for intensive-care prognosis have been based on the classic approach to logistic regression; however, this approach has its drawbacks. We describe these problems in the next section and investigate an alternative method based on Bayesian statistics.

## 2. BAYESIAN LOGISTIC REGRESSION

An assumption of *classic logistic regression* is that a conditional probability *p*(*y* = 1|**x**) is related to a vector of covariates **x** via a single model of the form

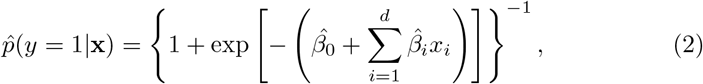

where 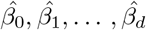 are the regression coefficients, and *x*_1_,…, *x*_*d*_ are the components of **x** (Multiplicative terms can be added to model interactions between explanatory variables). Furthermore, it is assumed that effective parameter values 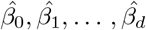 for this model can be obtained by *maximum-likelihood estimation*.

There are two problems with the classic approach. Firstly, there is the assumption that variables *y* and **x** are related by a single model with a prespecified structure *M*, but this does not take account of the fact that we are uncertain about *M*. Secondly, for a given choice of *M*, we are, in truth, uncertain about the vector ***β*** of parameter values associated with *M*, yet the maximum-likelihood approach imposes a single set of parameter values on *M*. In principle, these two criticisms can be addressed by regarding logistic regression within the framework of Bayesian statistics.^14,15^

*Bayesian statistics* provides a very different approach to the problem of unknown model parameters. Instead of considering just a single value for a model parameter, as done by maximum likelihood estimation, Bayesian inference expresses the uncertainty of parameters in terms of probability distributions and integrates them out of the distribution of interest.^16^ For example, by expressing the uncertainty in parameter vector ***β*** for a given model *M* as the posterior probability distribution *p*(***β***|*M*, D), where D is the observed data, we have

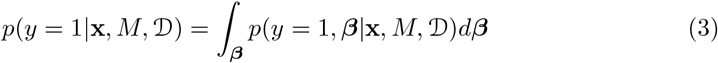

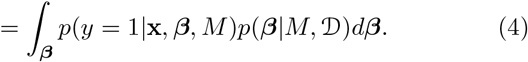

Where

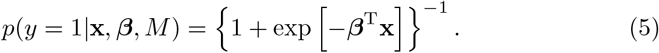

A common Bayesian assumption is that the posterior distribution *p*(*β*|*M*, D) is Gaussian, but, with *p*(*y* = 1| **x, *β***,*M*) defined by (4), the resulting integral in (4) cannot be solved analytically. However, Spiegelhalter & Lauritzen^17^ derived the approximation

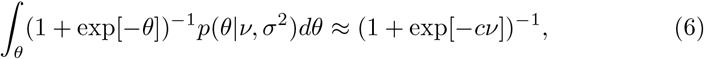

where *θ∼* Normal(*v, σ*^2^) and *c* is equal to (1 + *ξ*^2^*σ*^2^)^*−*1*/*2^ for an appropriate value of *ξ*. This was done via the approximation

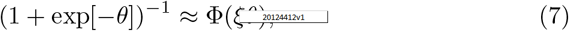

where Φ is the probit function.^18^ Because of the relationship between Φ and the error function, the latter can also provide an approximation for the left-hand side of (6).^19^

If *ξ*^2^ is set equal to *π/*8, as suggested by MacKay,^20^ then, from (5) and (6), we obtain the *Spiegelhalter–Lauritzen–MacKay (SLM) approximation*

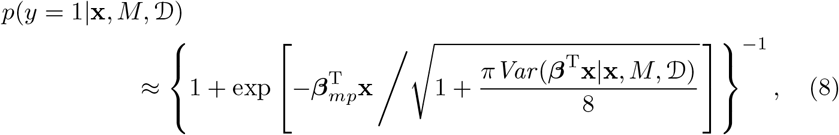

where ***β***_*mp*_ is the mode of *p*(***β***|*M, D*), and *Var* (***β***^T^**x**|**x**, *M, D*) is the variance of the posterior distribution *p*(***β***^T^**x** |**x**, *M*, D). Bishop discusses the error in ignoring the square root in (8).^21^

There is, however, still the uncertainty in the choice for model *M*. This uncertainty can be dealt with by averaging over all possible models:

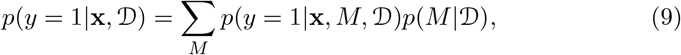

where *p*(*M*| *D*) is the posterior probability for model *M*. On substituting (4) into (9), we have the general expression for *Bayesian model-averaged logistic regression*:

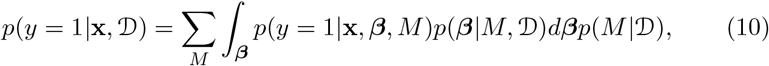

The SLM approximation provides an estimate of *p*(*y* = 1| **x**, *M*, D) for (9), but, in order to perform model-averaging, we also need the posterioir model probability *p*(*M*|D). In Section 4, we discuss the GLIB S-PLUS function for estimating this probability, but first we consider the evaluation of the mode and variance required for (8).

## 3. GIBBS SAMPLING

One route to estimating the mode and variance for (8) is to use the *evidence– framework* scheme proposed by MacKay^22^ and recommended by Bishop^21^; however, our experience (unpublished) has been that the approximations required to satisfy this scheme make it unreliable. Therefore, we obtained estimates of the regression coefficients via Gibbs sampling.

*Gibbs sampling* provides a Markov chain simulation of a random walk in the space of ***β***, which converges to a stationary distribution approximating the joint distribution *p*(***β***|*M, D*).^23^ In addition to providing an estimate of the mode ***β***_*mp*_, the stationary distribution also provides an estimate of the variance of ***β***^T^**x** via the estimated covariance for ***β***. The freeware BUGS package provides a convenient environment in which to conduct Gibbs sampling.^24^

## 4. GLIB

Bayesian model averaging can be performed using the freeware GLIB package,^25^ which is designed for use within S-PLUS.^26^ For a given set of models *M*_0_, *M*_1_,… *M*_*K*_ (where *M*_0_ is the null (intercept-only) model), GLIB can estimate the model posterior probabilities *p*(*M*_*k*_ |*D*) for each model through the use of Bayes factors. The *Bayes factor B*_*i*_,_*j*_ associated with models *M*_*i*_ and *M*_*j*_ is defined as

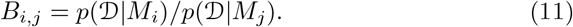

Raftery^27^ showed that application of the Laplace approximation^28^ to the integral in the expression

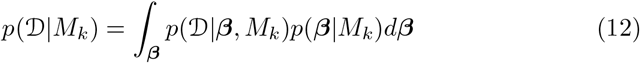

gives an approximation for ln *B*_*k*,0_ that can be calculated using quantities readily provided by regression packages such as GLIM.^29^ This enables *p*(*M*_*k*_|D) to be determined through the relationship

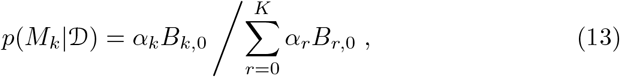

where *α*_*k*_ = *p*(*M*_*k*_)*/p*(*M*_0_). The prior distribution *p*(***β*** |*M*_*k*_) in (12) is defined by three hyperparameters: *v*_1_, *ψ*, and *φ*.^30^ GLIB fixes *v*_1_ and *ψ* to 1, and *φ* is set to 1.65 by default. Raftery & Richardson^31^ gave two examples illustrating the use of GLIB.

If the number of candidate models is very large, the total time taken by GLIB to compute *p*(*M*_*k*_|*D*) for each model can be lengthy. In such a situation, a pragmatic approach is to use the *bic*.*logit* S-PLUS function.^25^ This uses an approximation for ln *B*_*k*,0_^32^ based on the Bayesian Information Criterion.^33^ Although this approximation is less accurate than that used by GLIB for ln *B*_*k*,0_, *bic*.*logit* can filter out a large number of candidate models by implementing the following model-selection criteria.^34^

- **First criterion for model selection** If a model is far less likely a posteriori than the most likely model, it should be excluded. Therefore, exclude *M*_*k*_ if

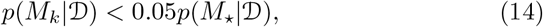

where *M*_*\**_ is the model with maximum *p*(*M*| *D*). This inequality is equivalent to

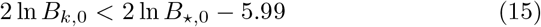

if *p*(*M*_*k*_) = *p*(*M*_*\**_) for all *k*.
- **Second criterion for model selection (Occam’s Window)** Exclude any model that receives less support from the data than a simpler model that is nested within it. Therefore, exclude *M*_*k*_ if there exists *M*_*j*_ nested within it for which

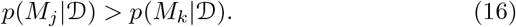

This inequality is equivalent to

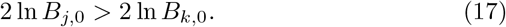

## 5. EXPERIMENTAL

### 5.1. Data

The 327 patients comprising the dataset were present in the adult ICU at St Thomas’ Hospital, London, from January 1997 to July 1997. The 11 attributes of the dataset are those listed in Table 1, and the values were recorded during the first 24-hours of each patient’s stay in ICU.

**Table 1:** The attributes of interest

| Attribute | Name | Data type | Levels |
| --- | --- | --- | --- |
| Age (years) | *Age* | Continuous | — |
| Artificial ventilation required | *Vent* | Nominal | “1” = true; “0” = false |
| Type of inotrope support | *Ino* | Ordinal | “0” = no intotropes; “1” = dopamine; “2” = adrenaline only; “3” = adrenaline plus other inotrope(s) |
| Serum bilirubin ($\mu$ mol/l) | *Bili* | Continuous | — |
| Acute renal failure | *ARF* | Nominal | “1” = true; “0” = false |
| 24-h urine volume (ml) | *UVol* | Ordinal | “0” = (0 - 50ml); “1” = (51 - 300ml) ; “2” = (> 300ml) |
| Surgical category | *Cat* | Nominal | “1” = elective (mostly cardiothoracic); “2” = emergency (medical patients); “3” = emergency (general surgery) |
| Creatinine ($\mu$ mol/l) | *Creat* | Continuous | — |
| Left ventricular intercept | *LVI* | Continuous | — |
| Glasgow Coma Score | *GCS* | Continuous<sup>a</sup> | — |
| Died whilst in hospital<sup>b</sup> | *Died* | Nominal | “1” = true; “0” = false |

<sup>a</sup>Levels are 3, 4, . . . , 15; therefore, the variable can be regarded as continuous
<sup>b</sup>The outcome variable

The dataset was incomplete. Of the 11*×* 327 cells of the dataset, 75 (2%) were empty, resulting in 67 (20%) incomplete rows. In the context of classic logistic regression, Lai^35^ found that imputing the incomplete cells of this dataset with class-conditional medians was as effective as using values derived by the EM algorithm^36^; therefore we used class-conditional median imputation. However, we deleted the three rows for which the outcome values were missing.

For this experiment, the continuous variables were neither discretized nor transformed in any way. The nominal and ordinal variables were replaced by binary dummy variables, which resulted in a total of 13 candidate explanatory variables.

### 5.2. Classic logistic regression

With the 13 candidate explanatory variables present, a main-effects logistic regression model was assessed using the leave-one-out version of cross-validation.^37^ The regression coefficients were obtained from the S-PLUS *glm* function, and the pooled predicted probabilities were assessed with respect to the corresponding (half) Brier score and ROC-plot area (Table 3). The Brier score measured predictive accuracy whereas the ROC-plot area measured discrimination.^37^

**Table 2:** Brier scores and ROC-plot areas (*A*_*z*_) resulting from the Bayesian and classical logistic regressions. The bootstrap 95% confidence intervals for the Brier scores and the estimated standard errors for the ROC-plot areas are also given.

| Type of logistic regression | Brier | 95%CI(Brier) | $A_z$ | $\widehat{SE}(A_z)$ |
| --- | --- | --- | --- | --- |
| Classical | 0.142 | (0.113, 0.170) | 0.826 | 0.026 |
| Bayesian with SLM approximation | 0.152 | (0.126, 0.182) | 0.821 | 0.026 |
| Bayesian without SLM approximation | 0.162 | (0.129, 0.198) | 0.815 | 0.027 |

**Table 3:** Brier scores and ROC-plot areas (*A*_*z*_) resulting from the use of single models and model averaging.

| Type of logistic regression | Brier | 95%CI(Brier) | $A_z$ | $\widehat{SE}(A_z)$ |
| --- | --- | --- | --- | --- |
| Single model with 13 variables | 0.133 | (0.110, 0.162) | 0.848 | 0.024 |
| Single model with 10 variables via stepwise selection | 0.130 | (0.106, 0.156) | 0.848 | 0.024 |
| Model averaging ($\phi = 1.65$) | 0.139 | (0.116, 0.164) | 0.820 | 0.027 |
| Model averaging ($\phi = 1$) | 0.139 | (0.118, 0.167) | 0.817 | 0.027 |
| Model averaging ($\phi = 5$) | 0.138 | (0.118, 0.167) | 0.820 | 0.027 |

In an effort to reduce the number of explanatory variables, stepwise variable selection was performed using the S-PLUS *step* function set at its default values, whereby variable selection was based on the Akaike Information Criterion.^38^ The resulting logistic regression model, which consisted of 10 explanatory variables, was evaluated by leave-one-out cross-validation (Table 3).

### 5.3. Bayesian logistic regression

Gibbs sampling was performed with BUGS (in the form of WinBUGS^39^) using 40-fold cross-validation. A non-informative prior for the regression coefficients was approximated by a normal distribution with mean 0 and variance 10^6^. Each Markov chain consisted of 11,000 samples, including an initial ‘burn-in’ of 1,000 samples.

In order to reduce correlations, and thus improve convergence, the covariates were reparameterized by centering them about their respective means.^40^ The improvements to convergence obtained by this reparameterization were confirmed by the Raftery-Lewis^41^ and Gelman-Rubin^42^ diagnostics provided by the freeware CODA diagnostic package.^43,44^

### 5.4. Results from Bayesian logistic regression

Although classical logistic regression gave better results than that obtained from Gibbs sampling with the SLM approximation (Table 2), the differences were not significantly different with respect to either Brier-score terms or ROC-plot (p-value *>* 0.15). However, omission of the SLM approximation did make a significant difference to the Brier-score terms (p-value = 0.008). Because of the lack of improvement when using Gibbs sampling, we decided to conduct the model-averaging phase of the study using maximum-likelihood estimates for the regression coefficients. In other words, (10) was replaced by

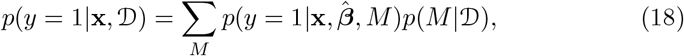

where 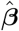 is the vector of regression coefficients estimated by maximum likelihood with respect to model *M*.

### 5.5. Bayesian model averaging

With 13 candidate explanatory variables, there were 2^13^ (i.e. 8192) possible models (including the null model). This was too many for GLIB to determine ln *B*_*k*,0_ for each model in a reasonable time; therefore, we initially used *bic*.*logit* to reduce the number of models to a more manageable subset. This produced a subset consisting of 40 models. Using the values for 2 ln *B*_*k*,0_ for these 40 models provided by GLIB, we applied the two model-selection criteria described in Section 4 to this subset. This resulted in the selection of six models when the hyperparameter *φ* was set to 1.65. GLIB was rerun on the six models to obtain their estimated posterior probabilities *p*(*M*_*k*_| *D*).

In order to ascertain the sensitivity of the results to choice of hyperparameter, the GLIB phase of the analysis was repeated using different values for *φ*. When *φ* was set to 1, the model-selection criteria produced 12 models; with *φ* = 5, three models were selected. All the models of this study are listed in Table 4. The three sets of models were evaluated by leave-one-out cross-validation (Table 3).

**Table 4:**
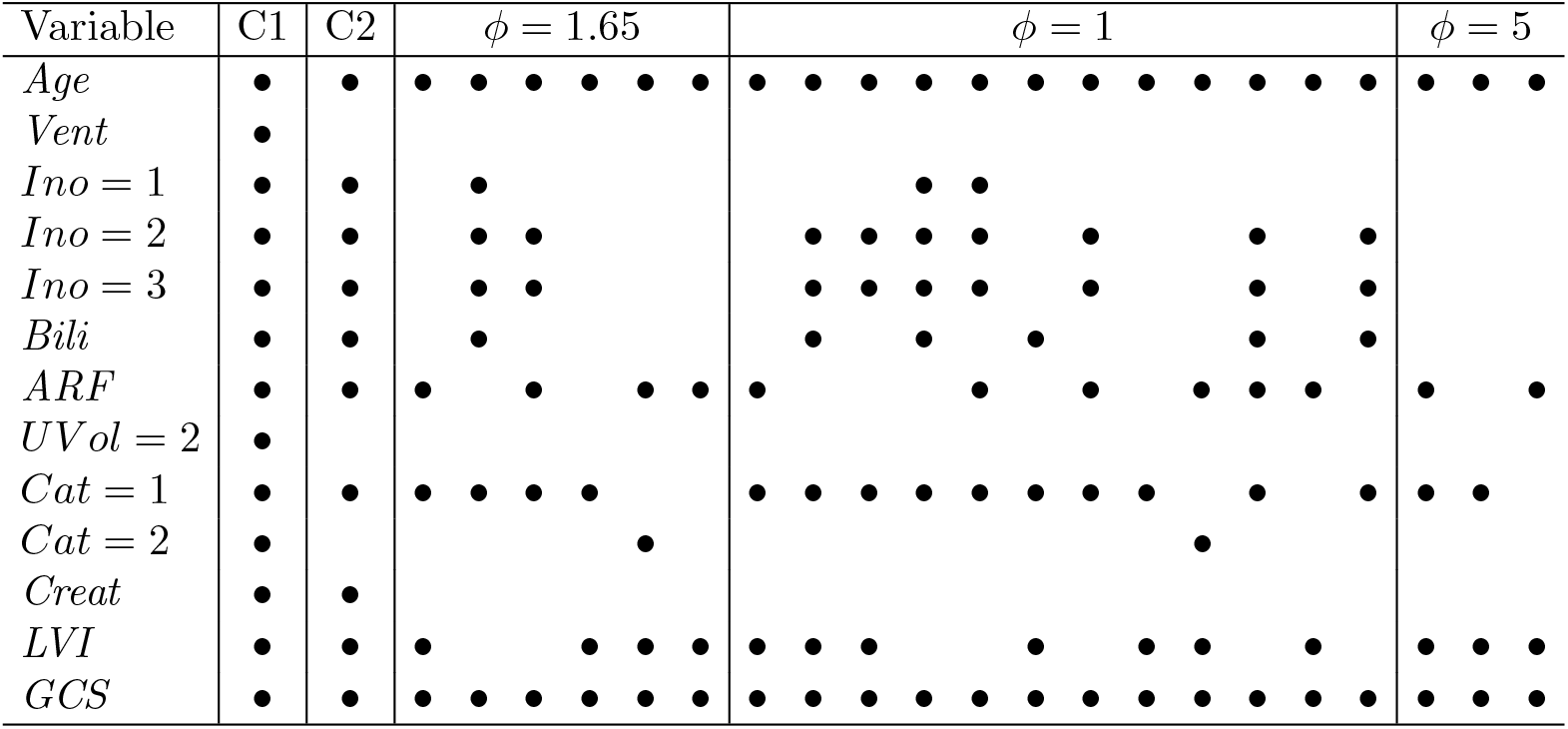
Models used in the study. C1 is the model containing all the candidate variables, and C2 is the model resulting from stepwise logistic regression. A black dot indicates that a variable was present. The 6, 12, and 3 models corresponding to *ϕ* = 1.65, *ϕ* = 1, and *ϕ* = 5, respectively, are listed.

### 5.6. Results from Bayesian model averaging

Table 3 gives the Brier scores and ROC-plot areas resulting from model averaging. Use of the single model with all the candidate variables present (Table 5) was significantly better than model averaging with respect to both Brier-score terms (p= 0.008) and ROC-plot area (p= 0.034).

**Table 5:** Regression analysis of the logistic regression model containing all the candidate variables according to maximum likelihood estimation. For each variable, the table gives the estimated regression coefficient (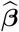), the estimated standard error for the regression coefficient 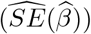, and the 95% confidence interval for the odds ratio (95%CI(OR)). The odds ratios for the continuous variables were calculated with respect to the first and third quartiles.

| *Variable* | $\hat{\beta}$ | $\widehat{SE}(\hat{\beta})$ | 95%CI(OR) |
| --- | --- | --- | --- |
| *(Intercept)* | -0.0431 | 1.37 |  |
| *Age* | 0.0599 | 0.0135 | (1.78, 4.44) |
| *Vent* | -0.382 | 0.548 | (0.23, 2.00) |
| *Ino* = 1 | 0.933 | 0.498 | (0.96, 6.75) |
| *Ino* = 2 | 1.593 | 0.500 | (1.85, 13.1) |
| *Ino* = 3 | 1.947 | 0.599 | (2.17, 22.6) |
| *Bili* | 0.00994 | 0.00508 | (1.00, 1.27) |
| *ARF* | 1.0910 | 0.588 | (0.94, 9.42) |
| *UVol* = 2 | -0.460 | 0.591 | (0.20, 2.01) |
| *Cat* = 1 | -1.375 | 0.514 | (0.09, 0.69) |
| *Cat* = 2 | 0.285 | 0.526 | (0.47, 3.73) |
| *Creat* | -0.00290 | 0.00156 | (0.62, 1.01) |
| *LVI* | -0.0322 | 0.0139 | (0.41, 0.93) |
| *GCS* | -0.266 | 0.0482 | (0.01, 0.13) |

## 6. DISCUSSION

In this paper, we have demonstrated that, in the context of ICU prognostic modelling, Bayesian logistic regression and Bayesian model averaging do not necessarily provide better predictive accuracy and discrimination than that given by a single regression model optimized by maximum likelihood estimation.

In the absence of any prior knowledge concerning the regression coefficients, we used a normal distribution with a very large variance to approximate a noninformative prior. However, if prior knowledge about some of the regression coefficients had been available to us (for example, from relevant publications), the Bayesian approach may have led to improved accuracy.

If we order the regression models in terms of their empirically-derived performance metrics then

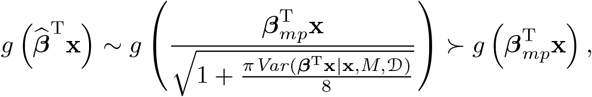

where *g*(*η*) = [1 + exp(*η*)]^*−*1^. But a curious aspect of this ordering is that we used a locally uniform prior, and from the Bayesian relationship

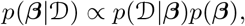

we would have expected the mode of the posterior distribution to virtually coincide with the maximum likelihood estimate 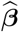. The reason for the observed ordering is not clear.

In spite of the claims made for the GLIB strategy,^31^ we did not find it to be superior to the classic, single-model approach. This may be due to the approximations for ln *B*_*k*,0_ being insufficiently accurate with respect to our dataset; however, with another ICU dataset, model-averaging may prove to be superior. Furthermore, model averaging has the advantage that it can exploit knowledge concerning the prior model probabilities *p*(*M*) used by (13).

In addition to the theoretical advantage to using model averaging (Section 2) there is also a disadvantage. With a single logistic regression model for probability *p*(*y* = 1| **x**), each regression coefficient (along with any associated multiplicative interaction terms) indicates the change in the probability for a unit change in the variable associated with the coefficient. Thus, the structure of the model provides some degree of interpretability. In model averaging, however, we are confronted with a collection of models, and if a number of models in the collection happen to have posterior probabilities close to that for the most probable model, model interpretation becomes much more complex.

## Data Availability

Data is from anonymised ICU patient data and is not available to the public.

## ACKNOWLEDGEMENT

We thank Dr David Treacher and Dr Alicia Vedio from the Intensive Care Unit at St Thomas’ Hospital, London, for permission to use the dataset.

